# The retirement-health puzzle: Breathe a sigh of relief at retirement?

**DOI:** 10.1101/2022.03.13.22271992

**Authors:** Shohei Okamoto, Erika Kobayashi

**Author notes:** Corresponding author: Dr Shohei Okamoto, Research Team for Social Participation and Community Health, Tokyo Metropolitan Institute of Gerontology: 35-2 Sakae-cho, Itabashi-ku, Tokyo, Japan 1730015. **Author contributions:** SO conceptualised the study, conducted the statistical analysis, and prepared a draft of the paper, which were refined by EK. All authors approved the final version of the paper and provide their consent for submission and publication.

## Abstract

**Objectives:** While the health effects of retirement have been well studied, existing findings remain inconclusive, and the mechanisms underlying the linkage between retirement and health are unclear. Thus, this study aimed to evaluate the effects of retirement on health and its potential mediators.

**Methods:** Using a national household survey conducted annually from 2004 to 2019 in Japan (the Japan Household Panel Survey), we evaluated the effects of retirement among Japanese men aged 50 or older on their health, in addition to other outcomes that could be attributed to health changes associated with retirement (i.e. health behaviours, psychological well-being, time use for unpaid activities, and leisure activities). As outcomes are not measured every year, we analysed 5,794–10,682 person-year observations by 975–1,469 unique individuals. To address the potential endogeneity of retirement, we adopted an instrumental variable fixed-effects approach based on policy changes in pension-eligible ages for employee pensions.

**Results:** We found that retirement improved psychological well-being, exercise habits, and time spent on unpaid work. The psychological benefits of retirement were no longer observed for longer durations after retirement, whereas healthy habits and unpaid activities continued. Moreover, health-related improvements after retirement occurred mostly in the higher-income group.

**Discussion:** Enhancement in personal quality of life owing to increased leisure time and stress reduction from work in addition to life style changes may be key to understanding the health benefits of retirement. Considering the mechanisms behind retirement–health relationships and potential heterogeneous effects is essential for healthy retirement lives when increasing the retirement age.

**Highlights:**

- In line with theories, previous studies report mixed results on effects of retirement on health.
- Empirical evidence on mechanisms underlying the linkage between retirement and health is scarce.
- Retirement effects on health and potential mediators are evaluated by a quasi-experimental approach.
- Retirement improves psychological well-being, exercise habits, time spent on unpaid work, and satisfaction with leisure.
- Health-related improvements after retirement occur mostly in the higher-income group.

## 1. **Introduction**

Industrialised societies emphasise delaying retirement timing in response to economic and financial challenges associated with population ageing. Researchers have been incrementally concerned about the effects of this policy change on the health of older populations; however, empirical studies have so far presented inconclusive findings on the health effects of retirement. The mechanisms underlying the association between retirement and health also remain unclear. Therefore, accumulating evidence on the health effects of retirement as well as uncovering potential channels that mediate the retirement-and-health relationship is crucially important in creating plausible environments for employment in later life, which would continue to be in demand in the context of population ageing.

### 1.1. Theoretical background

To understand the linkage between retirement (or employment) and health in later life, several theories have been applied to different areas of expertise. One of the most common theories used to conceptualise the relationship between retirement and health is the human capital model (i.e. the Grossman model) (Grossman, 2000). In the Grossman model, individuals maximise their utility, which is determined by their own health status and consumption of normal goods/services within budget and time constraints. Based on the Grossman model, retirement can have both protective and detrimental effects on one’s health. Individuals’ health may improve after retirement because retirement decreases the opportunity cost of health investments and increases the available time for health-producing activities. However, individuals may lose incentives to invest in their health after retirement, as their income is no longer associated with their health status, and they may face tighter budget constraints. Thus, the effect of retirement on health is ambiguous. From the Grossman model’s perspective, behavioural changes, including health behaviours, leisure activities, and consumption, are key to understanding the mechanisms of the health effects of retirement. Although non-health-related consumption may be viewed as a substitution of health, it is important to note that the pathway that even consumption of normal goods/services affects one’s health status can exist.

Although retirement itself may be viewed as a stressful life event (Minkler, 1981), it can also offer relief from a stressful working life (Bossé et al., 1991). Stress affects mental health and subjective well-being, which is also linked with physical health (Cross et al., 2018) due to behavioural changes, such as smoking, drinking, eating, and sleeping.

Employment may also generate latent consequences beyond its economic effects, including the psychosocial meaning of employment, such as a sense of purpose and goals, identity, and social contacts (Jahoda, 1981). Individuals may undergo changes in psychosocial aspects of their life upon retirement, which are important determinants of health (House et al., 1988; Windsor et al., 2015). However, the effects of retirement on psychological meaning and social relationships are ambiguous because retirees may obtain new roles and/or richer social networks with friends and community members with more leisure time available, even though they may lose job-related ones.

Thus, the overall effect of retirement on health is not explicit from these theories, with effects in both the negative and positive directions.

### 1.2. Literature review

To evaluate the causal effects of retirement on health and other health-related outcomes beyond mere association, it is essential to address the endogenous relationship between health and retirement. Most of the previous studies discussed below adopt quasi-experimental approaches to identify the effects of retirement on health and health behaviours, utilising policy changes (e.g. retirement age and state pension eligibility age). In alignment with the theories, findings from empirical studies remain inconclusive, with mixed results for the causal effects of retirement (or employment) on health in later life.

Many previous studies show that physical and/or mental health, which is measured by subjective or objective scales, improves after retirement, or deteriorates by staying in employment for longer durations (Atalay & Barrett, 2014; Coe & Zamarro, 2011; Eibich, 2015; Gorry et al., 2018; Heller-Sahlgren, 2017; Hessel, 2016; Kolodziej & Garcia-Gomez, 2019; Nishimura et al., 2018; Oshio & Kan, 2017; Rose, 2020; Shai, 2018). Other studies have also found negative effects of retirement on cognitive functioning (Bonsang et al., 2012; Celidoni et al., 2017; Kajitani et al., 2016; Rohwedder & Willis, 2010).

To help understand the link between retirement and health, existing studies suggest that improvements in health behaviours, such as physical activities, sleep duration, and smoking, are observed after retirement (Eibich, 2015; Kampfen & Maurer, 2016; Kesavayuth et al., 2018). Similarly, positive health effects and behavioural changes in the utilisation of healthcare services after retirement have been reported (Frimmel & Pruckner, 2020).

In contrast, studies so far have found contradictory or nonsignificant results. Several studies observe worsened physical and/or mental health owing to retirement or positive health effects of employment in later life (Behncke, 2012; Calvo et al., 2013; Okamoto et al., 2018) as well as increased mortality after retirement (Fitzpatrick & Moore, 2018). Other studies have found weight gain (Chung et al., 2009; Godard, 2016) and increased healthcare use after retirement (Lucifora & Vigani, 2018; Zhang et al., 2018). Similarly, by a longer working horizon, a previous study reports increased exercise and positive effects on obesity and self-reported satisfaction with health (Bertoni et al., 2018).

Other studies find no evidence that retirement or reform to increase the normal retirement age affects cognition, mortality, or healthcare utilisation (Hagen, 2017; Hernaes et al., 2013; Rose, 2020), as well as no immediate effects of retirement on behavioural outcomes (Rose, 2020).

Even with these mixed findings, the drivers of the differences are not well explored, and can be driven by regional or cohort heterogeneity as well as methodological differences, such as outcome measures and identification strategies. Therefore, much evidence needs to be accumulated to better understand the association between retirement and health. In addition, with the scarce empirical evidence for the mechanisms by which retirement affects health, it is necessary to assess the effects of retirement on a wide variety of outcomes that can mediate the linkage between retirement and health. Although retirement effects on health behaviours have been investigated so far to unveil the underlying mechanisms, there can be other factors unexplored as retirement is a big life event that induces various changes in one’s life.

### 1.3. Summary of this study

This study aimed to assess the effect of retirement on health as well as a wide variety of outcomes that can mediate the linkage between retirement and health, including health behaviours, subjective well-being, time spent on unpaid work, and consumption composition. To do so, we analysed the data derived from a nationally representative sample of Japanese adults using an instrumental variable (IV) approach, and by policy changes in the public pension eligibility age for employee pensions. Our study expands the literature in the field by evaluating the effect of retirement on a wide variety of outcomes, which could help understand the mechanisms of the linkage between retirement and health.

Analysis using an IV fixed-effects model resulted in a main finding which suggests that retirement leads to an enhancement in psychological distress and subjective well-being as well as increases in exercise habits and time spent on unpaid work, whereas the effects on health measured by self-rated health and body mass index were not observed. Additional analysis suggests that the psychological benefits of retirement may diminish over longer durations after retirement. Furthermore, by subgroup analysis, we observed the heterogeneous effects of retirement, particularly by income, finding that the improvements in dependent variables after retirement occurred mostly among the higher-income group, but not in the lower-income group.

The remainder of this paper is organised as follows. The next section presents the institutional setting in Japan regarding retirement policies and the public pension system. In the following sections, methods of the study that include descriptions of the data and variables are introduced, followed by a section to explain the identification strategy. Subsequently, we present the results and discuss our findings, as well as their limitations, followed by conclusions.

## 2. Institutional setting

### 2.1. Retirement policy in Japan

Under the *Act on Stabilization of Employment of Elderly Persons*, enacted in 1986 (its origin was enacted in 1971), the minimum retirement age has been amended to secure the employment of older populations, corresponding to an increase in pensionable age for employee pensions. In 1998, the amendment of the Act was enforced, thereby mandating the minimum mandatory retirement age to be 60 or later for employers that adopted the mandatory retirement scheme. From 2006, by the 2004 amendment of the Act, for employers that set their mandatory retirement age below 65, any one of three schemes were required: (1) raise the mandatory retirement age to 65 or later; (2) abolish the mandatory retirement scheme; and (3) introduce a continuous employment system until the age of 65. The minimum age required for these schemes by low gradually increased from 60 to 65 years until 2013. Before the enforcement of the amendment in 2012, employers were allowed to have a labour-management agreement that defined non-eligible workers for the continuous employment system; however, this exceptional measure was abolished in 2013 and, in principle, all workers who wish to be continuously employed until the age of 65 have become eligible for the system. With the amendment enforced in 2021, employers that set their retirement age to be between 65 and 69 or the Continuous Employment System until the age of 65 have been obligated to make efforts to secure employment until the age of 70.

### 2.2. The national pension system

The old-age public pension in Japan, which is a pay-as-you-go universal pension coverage in which residents in Japan aged between 20 and 60 make contributions, consists of two main elements: basic pension and employee pension. To be concrete, there are three main types of individuals assumed in this pension scheme: (Category 1) those only enrolled in the basic pension scheme, paying flat-amount pension contributions (e.g. self-employed individuals), and who will receive flat-rate old-age pension benefits; (Category 2) those enrolled in employees’ pension scheme in addition to the basic pension scheme, bearing contributions at a certain proportion of their earnings (around 18%) with the half paid by employers, and who will receive earning-related pension benefits (e.g. employees and public officials); (Category 3) those who do not bear pension contributions by themselves and who will receive flat-rate pension benefits (e.g. dependents of category 2 workers). Among all the insured individuals in the public pension scheme, the largest proportion falls in the category of category 2 workers (around 66%) (Ministry of Health, Labour and Welfare, 2021).

For employee pensions, based on the year and month of birth and sex, the default pensionable age is raised from 60 to 65 between 2001 and 2013 for the flat-rate pension and between 2013 and 2025 for the earning-related pension; for women, they are to be raised five years behind schedules for the men’s pension reforms.

## 3. Methods

### 3.1. Data

The data for this study were obtained from the Japan Household Panel Survey (JHPS/KHPS), which comprises a nationwide sample of Japanese aged 20 or over. The sample was extracted by a two-stage stratified random sampling method, using 24 levels according to regional and city classifications throughout Japan as the first stage and basic resident registers as the second stage. The survey began in 2004 and is conducted annually; new samples were added using the same procedure in 2007, 2009, and 2012. The website of the JHPS/KHPS provides information in greater detail (Panel Data Research Center, 2021).

To evaluate the effect of retirement on one’s later life, we only analysed individuals aged 50 years or over in this study. We restricted our analyses to male participants, since the retirement decision making of women is affected by various factors (e.g. marriage and informal caregiving) than that of men (Yamada & Shimizutani, 2015), which makes identification difficult. Moreover, we did not include self-employed individuals in our study because they usually receive a different type of public pension from employees (e.g. *National Pension*), and their retirement process tends to be unlike employees. Calculation of the final sample size is described below.

### 3.2. Retirement

In studies on retirement, the definition of retirement is important. Following the idea suggested by Lazear (1986) that retirement refers to being out of the labour force with no intent to seek employment, we identified retired individuals based on their labour force status, defining those not in work for pay at the time of the survey as retired. To isolate the effects of retirement from an unusual labour force status, such as unemployment and temporal job leave, we excluded those seeking a job or suspending from the job during the past year. Moreover, we restricted the sample to those whose retirement status changed during the study period in order to estimate the effects of the status change on dependent variables.

### 3.3. Dependent variables

To understand the effect of retirement on health and other potential mediating factors, the following self-reported dependent variables were analysed: As not all dependent variables were asked in every wave, we conducted available case analyses for each dependent variable. In Appendix Table A-1, we summarise the years of surveys in which each dependent variable is included.

#### 3.2.1. Subjective health and psychological well-being

While the JHPS/KHPS only includes a limited number of health outcomes, we use self-rated health (SRH) and psychological distress in addition to domain-specific and life satisfaction as indicators of subjective health and psychological well-being.

SRH is a five-point Likert scale question about one’s overall health status, which is widely used as a useful predictor of clinical outcomes (Fayers & Sprangers, 2002). We dichotomised SRH to take one if individuals reported bad or very bad health status, and zero otherwise.

Changes in psychological factors such as subjective well-being after retirement may mediate the linkage between retirement and health, as it is reported to be associated with health status (Diener & Chan, 2011)

Psychosomatic symptoms, which reflect one’s perceived physical and mental health status that can be related to depression (Kapfhammer, 2006), are defined as a summary score of frequencies (i.e. 1: never - 4: often) for nine questions about anxiety and some symptoms: (a) headache and dizziness, (b) palpitation and breathlessness, (c) poor digestion, (d) shoulder pain/backache, (e) easily tired, (f) easily catch a cold, (g) troublesome to meet people, (h) anxious about one’s current life, and (i) anxious about one’s future life. We standardised the score to take values between 0 and 1, with a higher value indicating better conditions, after reversing the scaling and adding up valid responses to each item. We further conducted analysis, decomposing the indicator into two scores: somatic symptoms comprising the former six items and psychological distress comprising the latter three items.

Another source of psychological well-being measure is one’s satisfaction with one’s life in general, scoring 0–10 with a higher value indicating higher satisfaction.

#### 3.3.2 Physical health

To measure one’s physical health status, we defined, using body mass index (BMI; see below for its definition), individuals with a BMI of 25.0≤BMI as overweight and a BMI 30.0≤BMI as obese.

In instances where individuals received a health check-up during the past year, they were asked if any health issues were found as a result of the check-ups. We used eight items as indicators of physical health: blood pressure, bone density, heart, anaemia, liver, kidney, diabetes, and metabolism.

#### 3.3.3. Health behaviours and healthcare utilisation

Health behavioural change is key to associating retirement with health (Eibich, 2015). As indicators of health behaviours, which can mediate the linkage between retirement and health, we analysed five variables: BMI, cigarette smoking, alcohol consumption, hours of sleep, exercise.

BMI, which is a measure of one’s nutritional status, was calculated as body weight (kg) divided by the square of the body height (m). As body height and body weight, obtained as self-reported measures, were not frequently asked in the survey, both measures were imputed by linear interpolation.

Smoking and alcohol consumption were dichotomised to indicate current smokers and those who drink alcoholic beverages more than once a week. Hours of sleep count the weekdays and weekends average, excluding the top/bottom 1% values as outliers. Exercise was measured as days of non-work-related exercise per week.

Moreover, we evaluate the retirement effects on healthcare utilisation, measured by health check-up, total health expenditure, and healthcare utilisation. Health check-up was dichotomised to indicate those who received a health check-up (i.e. periodic health examination by insurers or full medical check-up) during the past year. Total health expenditure was defined as a proportion of overall health spending among one’s entire consumption in the last month (See *leisure activities* below for detailed explanations). Healthcare use is a binary variable that takes the value of one if a respondent used outpatient or inpatient healthcare services or purchased over-the-counter drugs during the past year.

#### 3.3.4. Time-use for unpaid work

Considering that retirees may spend their time differently before retirement, changes in time use induced by retirement can be mediators of the association between retirement and health, as these activities may also provide latent effects, similar to paid jobs. As the JHPS/KHPS has continuously asked the participants for their per-day hours of unpaid work (i.e. volunteer, domestic work, and childcare), we assessed the effects of retirement on these variables. For childcare, we restricted the sample to those reporting to have a child or grandchild. To deal with outliers, we excluded the top 1% of these variables.

#### 3.3.5. Leisure activities

Some categories of leisure entertaining activities, likewise time use for unpaid work, can affect one’s health status directly or indirectly through purchases of goods and services. With an argument of the retirement-consumption puzzle, retirees may spend less on their work-related expenditures whilst sustaining or increasing other categories of expenditures (Hurst, 2008). We used consumption in the last month of the survey for eating out, culture and recreation, and social expenses (Also, this procedure is applicable for total health spending). To avoid evaluating the effects of an income decline associated with retirement, rather than a status shift and changes in their time allocation from work to retirement, we used consumption compositions, defined by the proportion of each expenditure among the total consumption. We excluded the top 1% values as outliers because some individuals reported unreasonable answers (e.g. the expenditure for entertainment exceeded total expenditure).

In addition to consumption, we used one’s satisfaction with leisure quantity (i.e. length) and quality (i.e. how to spend), scoring 0–10 with a higher value indicating higher satisfaction.

### 3.4. Empirical strategy

One of the key challenges in evaluating the health effects of retirement is that retirement is not randomly assigned. The standard ordinary least squares regression provides unbiased estimates with the orthogonality condition satisfied. However, in the topic of the retirement-and-health relationship, this is unlikely to be true, because retirement is a self-determined behaviour, which is affected by one’s health status and other unobserved factors; hence, endogeneity bias needs to be addressed by an econometric specification. Following the IV approach adopted by many previous works discussed earlier, and by using policy changes in public pension eligibility age, the association between the dependent variables of interest and retirement is formalised as

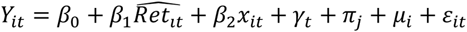

where *Y_it_* denotes the dependent variables, 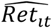 is the predicted retirement probability from the first stage for individual i in year t, and *x_it_* is a vector of control variables. Because variables that are largely changed by retirement (e.g. income) are ‘bad controls’ (Angrist & Pischke, 2009), we controlled for age, age squared, marital status (single or married), living alone, and home ownership. To obtain the predicted value for retirement from the first stage, we estimate the following equation:

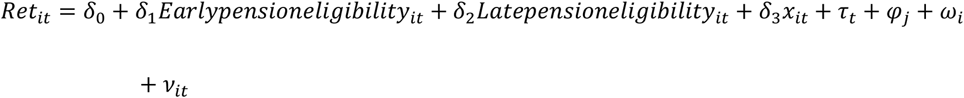

where one’s retirement status is predicted by two dichotomised instruments of the Employees’ Pension eligibility for the flat benefits (*early pension eligibility*) and the wage-proportional benefits (*late pension eligibility*). To additionally control for unobserved factors, we included year fixed effects (*γ_t_* and *τ_t_*), prefecture-by-scale fixed effects (*π_j_* and *φ_j_*), and individual fixed effects (*μ_i_* and *ω_i_*). Idiosyncratic errors in the second and first stages are expressed as *ε_it_* and *ν_it_*, respectively. Intuitively, these fixed effects control for the effects of each year common across all prefectures (e.g. business cycle in the whole country), effects of each prefecture by scale common across time (e.g. industrial structures of the place in a short period), and time-invariant individual characteristics (e.g. gene). Based on the Frisch–Waugh–Lovell theorem, prefecture-by-scale fixed effects were treated as partialling-out exogenous regressors to address the issue that the covariance matrix of orthogonality conditions is not of full rank (Angrist & Pischke, 2009).

For instruments to be valid, they need to meet the relevance and exclusion restrictions. First, pension eligibility is required to predict one’s retirement, which is empirically testable. Second, to meet the exclusion restriction, pension eligibility needs to affect the dependent variable solely through retirement. The pensionable age is determined by the government; thus, pension eligibility is assumed to be exogenous for individuals.

To estimate the association between retirement and health and other variables, we fitted a linear model (or a linear probability model for binary dependent variables). For the dependent variables of exercise days, time use for unpaid work, and proportions of each expenditure, the distributions were skewed, including many zero observations. To avoid model misspecification, these variables were approximated to the natural logarithm by inverse hyperbolic sine transformation (Burbidge et al., 1988).

We excluded those with missing independent variables because only a small number of individuals lacked the information (approximately 1% of the target sample). Thus, we used a pairwise deletion, avoiding unnecessary noises by imputing dependent variables (von Hippel, 2016). Because of the nature of a fixed-effects model, singleton observations were not used for the parameter estimates. Consequently, the final sample size comprises 5,794– 10,682 person-year observations by 975–1,469 unique individuals. Meanwhile, in a panel data analysis, potential biases because of sample attrition cannot be ignored. To partially address a non-response bias, all analyses were weighted by longitudinal weights, estimated by logit models as inverse probabilities of responding to each subsequent wave, conditional on respondents’ age, sex, marital status, education, employment status, and residential area at baseline.

All analyses were conducted using Stata MP, version 17.0, (StataCorp LLC, College Station, USA). Ethical approval was not required, as this study comprised a secondary analysis of publicly available data.

## 4. Results

### 4.1. Descriptive statistics

Table 1 reports descriptive statistics of dependent and independent variables in addition to instruments, divided by retirement status. Retirees reported bad health more than non-retirees (23.0% and 13.4%), whereas proportions of those being obese, overweight, smoking, and consuming alcoholic beverages were smaller in retirees. For most of the physical health issues, retirees had worse health status than non-retirees, which may be due to age differences between these groups. Retirees had longer hours of sleep, more frequent exercises, and used more healthcare services. On average, retirees were more satisfied with their leisure, health, and life than non-retirees. Except for childcare, retirees spent longer time on non-paid productive activities.

**Table 1.**
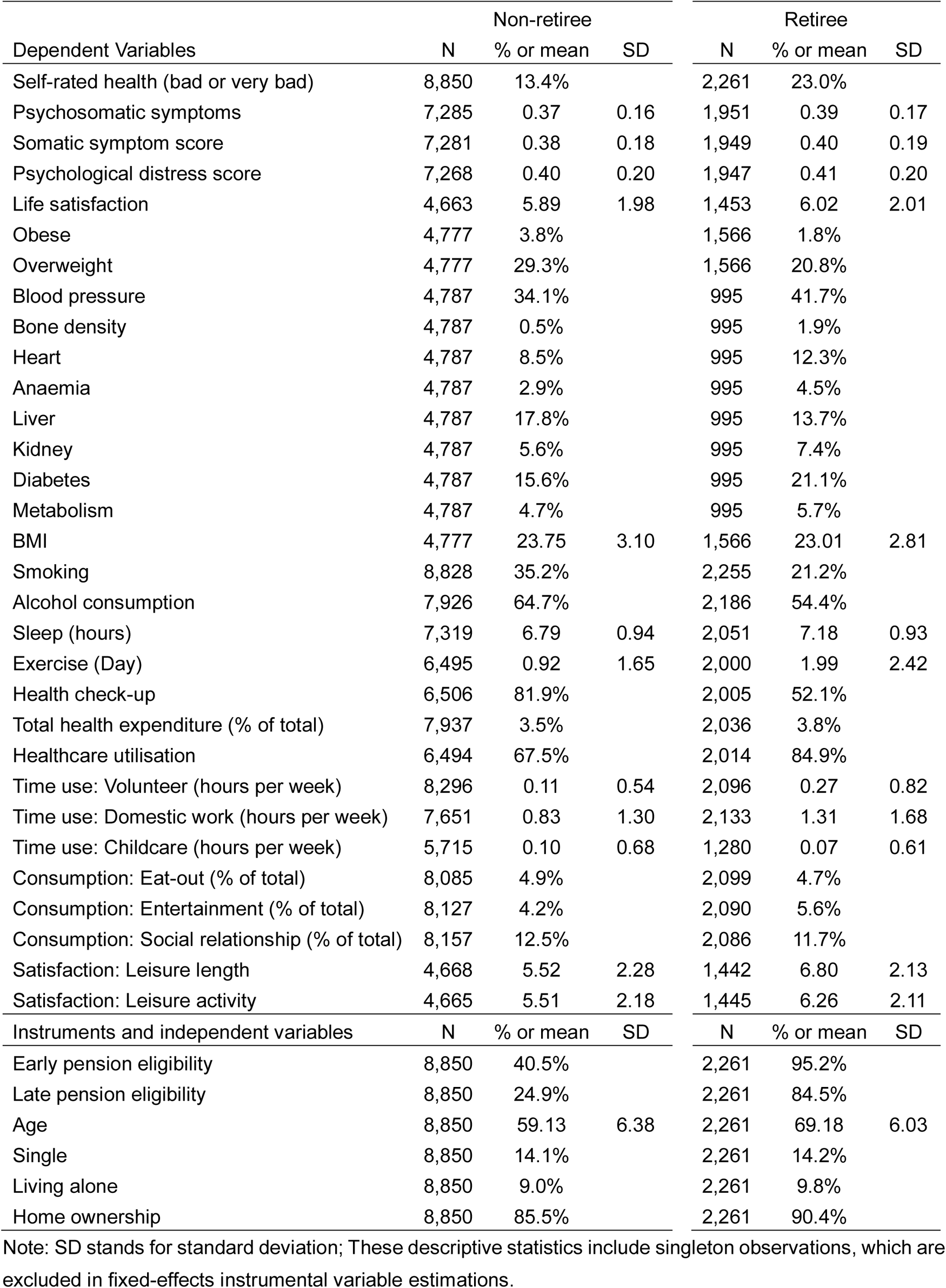
Descriptive statistics

| Dependent Variables | Non-retiree |  |  | Retiree |  |  |
| --- | --- | --- | --- | --- | --- | --- |
|  | N | % or mean | SD | N | % or mean | SD |
| Self-rated health (bad or very bad) | 8,850 | 13.4% |  | 2,261 | 23.0% |  |
| Psychosomatic symptoms | 7,285 | 0.37 | 0.16 | 1,951 | 0.39 | 0.17 |
| Somatic symptom score | 7,281 | 0.38 | 0.18 | 1,949 | 0.40 | 0.19 |
| Psychological distress score | 7,268 | 0.40 | 0.20 | 1,947 | 0.41 | 0.20 |
| Life satisfaction | 4,663 | 5.89 | 1.98 | 1,453 | 6.02 | 2.01 |
| Obese | 4,777 | 3.8% |  | 1,566 | 1.8% |  |
| Overweight | 4,777 | 29.3% |  | 1,566 | 20.8% |  |
| Blood pressure | 4,787 | 34.1% |  | 995 | 41.7% |  |
| Bone density | 4,787 | 0.5% |  | 995 | 1.9% |  |
| Heart | 4,787 | 8.5% |  | 995 | 12.3% |  |
| Anaemia | 4,787 | 2.9% |  | 995 | 4.5% |  |
| Liver | 4,787 | 17.8% |  | 995 | 13.7% |  |
| Kidney | 4,787 | 5.6% |  | 995 | 7.4% |  |
| Diabetes | 4,787 | 15.6% |  | 995 | 21.1% |  |
| Metabolism | 4,787 | 4.7% |  | 995 | 5.7% |  |
| BMI | 4,777 | 23.75 | 3.10 | 1,566 | 23.01 | 2.81 |
| Smoking | 8,828 | 35.2% |  | 2,255 | 21.2% |  |
| Alcohol consumption | 7,926 | 64.7% |  | 2,186 | 54.4% |  |
| Sleep (hours) | 7,319 | 6.79 | 0.94 | 2,051 | 7.18 | 0.93 |
| Exercise (Day) | 6,495 | 0.92 | 1.65 | 2,000 | 1.99 | 2.42 |
| Health check-up | 6,506 | 81.9% |  | 2,005 | 52.1% |  |
| Total health expenditure (% of total) | 7,937 | 3.5% |  | 2,036 | 3.8% |  |
| Healthcare utilisation | 6,494 | 67.5% |  | 2,014 | 84.9% |  |
| Time use: Volunteer (hours per week) | 8,296 | 0.11 | 0.54 | 2,096 | 0.27 | 0.82 |
| Time use: Domestic work (hours per week) | 7,651 | 0.83 | 1.30 | 2,133 | 1.31 | 1.68 |
| Time use: Childcare (hours per week) | 5,715 | 0.10 | 0.68 | 1,280 | 0.07 | 0.61 |
| Consumption: Eat-out (% of total) | 8,085 | 4.9% |  | 2,099 | 4.7% |  |
| Consumption: Entertainment (% of total) | 8,127 | 4.2% |  | 2,090 | 5.6% |  |
| Consumption: Social relationship (% of total) | 8,157 | 12.5% |  | 2,086 | 11.7% |  |
| Satisfaction: Leisure length | 4,668 | 5.52 | 2.28 | 1,442 | 6.80 | 2.13 |
| Satisfaction: Leisure activity | 4,665 | 5.51 | 2.18 | 1,445 | 6.26 | 2.11 |
| Instruments and independent variables | N | % or mean | SD | N | % or mean | SD |
| Early pension eligibility | 8,850 | 40.5% |  | 2,261 | 95.2% |  |
| Late pension eligibility | 8,850 | 24.9% |  | 2,261 | 84.5% |  |
| Age | 8,850 | 59.13 | 6.38 | 2,261 | 69.18 | 6.03 |
| Single | 8,850 | 14.1% |  | 2,261 | 14.2% |  |
| Living alone | 8,850 | 9.0% |  | 2,261 | 9.8% |  |
| Home ownership | 8,850 | 85.5% |  | 2,261 | 90.4% |  |

Note: SD stands for standard deviation; These descriptive statistics include singleton observations, which are excluded in fixed-effects instrumental variable estimations.

### 4.2. IV results

In Table 2, we present the first-stage results of an IV estimation. We confirmed that our instruments of two types of pension eligibilities accurately predict one’s retirement (Early pension, b: 0.06, robust standard error [SE]: 0,01, late pension, b: 0.11, and SE: 0.02, Cragg-Donald Wald F-statistic: 61.34). Figure 1 and 2 shows the estimated results for the health effects of retirement by IV fixed-effects regression in addition to the estimated effects of retirement on dependent variables, which can mediate the linkage between retirement and health, are presented. The full results for Figures 1 and 2 are presented in Appendix Tables A-2, A-3, A-4, and A-5.

**Figure 1.**
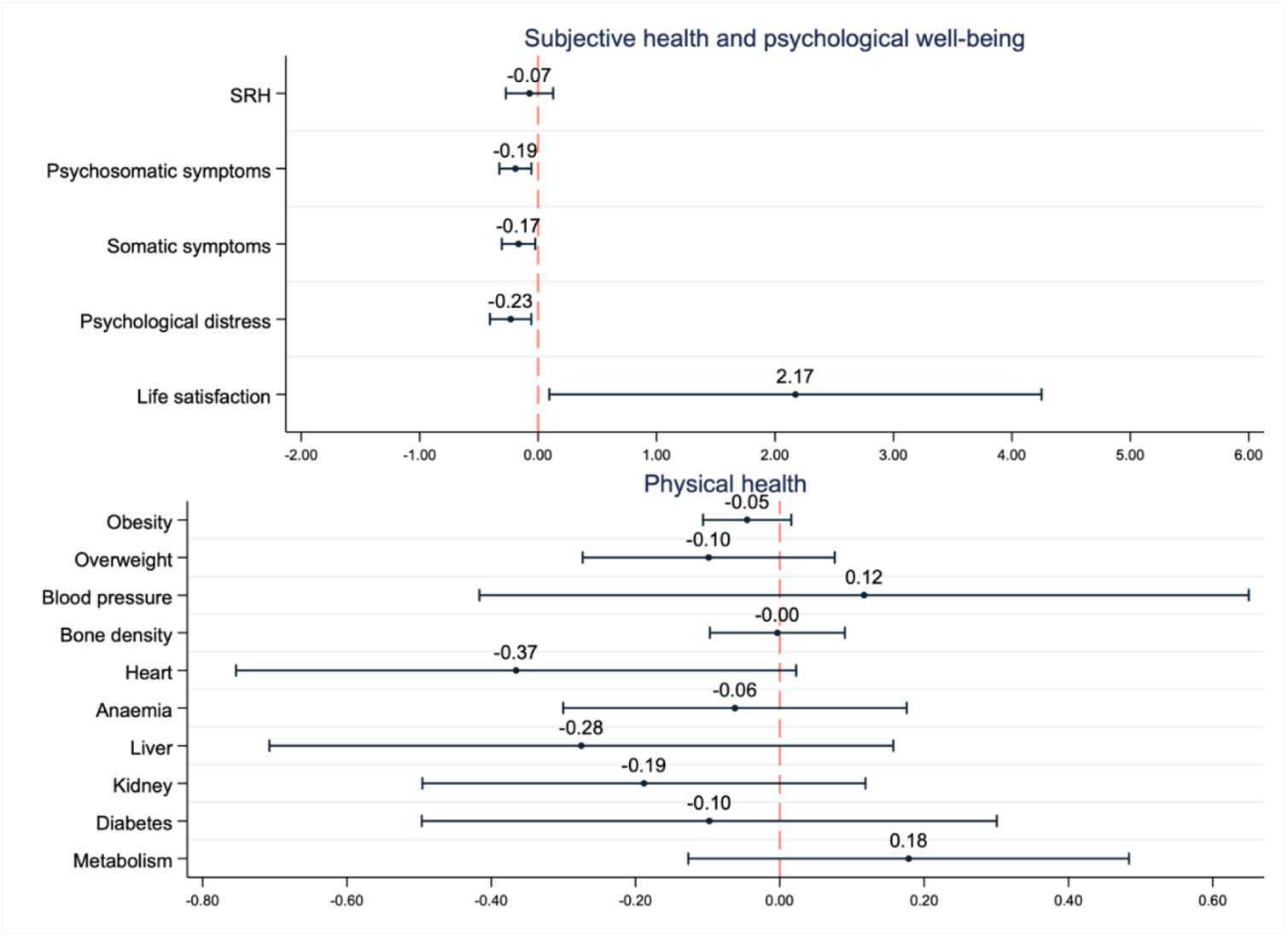
Impacts of retirement on health and psychological well-being Note) Plots are estimated coefficients with bars representing 95% confidence intervals from robust standard errors, obtained as the second stage results of IV estimations; All models control for age, age-squared, marital status (single or not), living alone, home ownership, individual-fixed-effects, year-fixed-effects, and prefecture-by-scale-fixed-effects; Prefecture-by-scale dummy is partialled out from all the other variables, using the Stata’s *partial* option; Singleton groups are not used for estimations; Full results are presented in Appendix Table A-2 and A-3.

**Figure 2.**
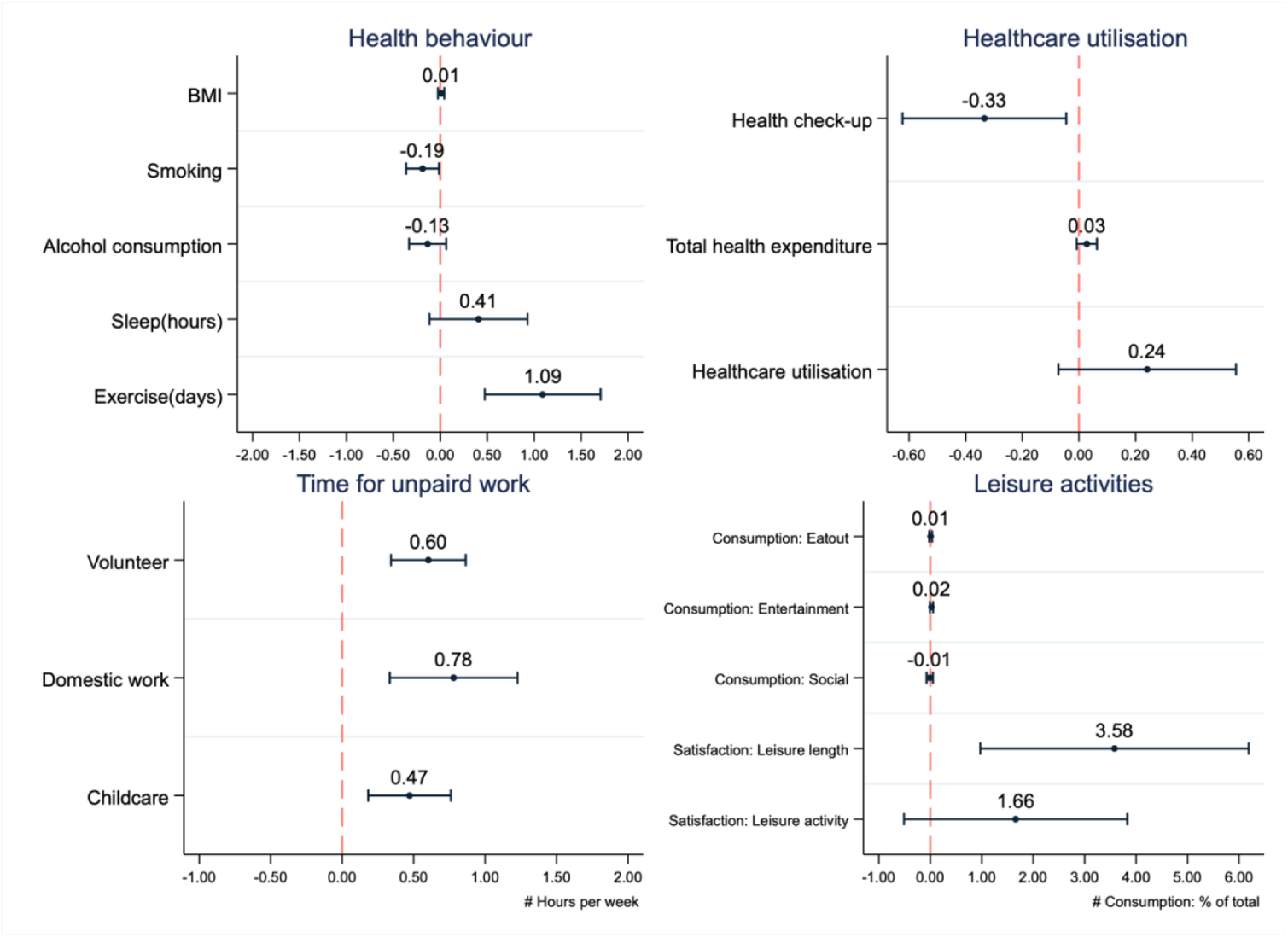
Impacts of retirement on health behaviours, time use, and leisure activities Note) Plots are estimated coefficients with bars representing 95% confidence intervals from robust standard errors, obtained as the second stage results of IV estimations; All models control for age, age-squared, marital status (single or not), living alone, home ownership, individual-fixed-effects, year-fixed-effects, and prefecture-by-scale-fixed-effects; Prefecture-by-scale dummy is partialled out from all the other variables, using the Stata’s *partial* option; Singleton groups are not used for estimations; Full results are presented in Appendix Table A-4 and A-5.

**Table 2.** First-stage result

| Retirement |  |
| --- | --- |
| Early pension eligibility | 0.06** (0.01) |
| Late pension eligibility | 0.11** (0.02) |
| Age | -0.17** (0.01) |
| Age<sup>2</sup>/100 | 0.16** (0.01) |
| Single | -0.03 (0.03) |
| Living alone | 0.05* (0.02) |
| Home ownership | 0.09** (0.03) |
| Individual FE | Yes |
| Year FE | Yes |
| Prefecture-by-scale FE | Yes |
| Cragg-Donald Wald F statistic: | 61.34 |
| Hansen J statistic (p-value) | 0.67 |
| Observations | 1,469 |
| Individuals | 10,682 |

Note: The dependent variable of the second stage estimation is self-rated health; Values are coefficients with robust standard errors in parenthesis; Singleton groups are not used for estimations; Prefecture-by-scale dummy is partialled out from all the other variables, using the Stata's *partial* option; FE denotes fixed-effects.

### 4.3. Effects on health and psychological well-being

Among the health measures, we found that retirement was associated with psychological well-being, showing a decline in psychosomatic symptoms (b: −0.19, SE: 0.07), somatic symptoms (b: −0.17, SE: 0.07), and psychological distress (b: −0.23, SE: 0.09) in addition to improvements in one’s life satisfaction (b: 2.17, SE: 1.06). Meanwhile, post-retirement changes in other outcomes of SRH and physical health status were not found to be significant.

### 4.4. Health behaviours and healthcare utilisation

We found a decline in smoking probability (b: −0.19, SE: 0.09) and an increase in exercise days (b: 1.09, SE: 0.31) after retirement. Notably, a drop in the probability of receiving health check-ups was observed after retirement (b: −0.33, SE: 0.15). Meanwhile, we did not observe significant post-retirement changes in alcohol consumption, hours of sleep, total health expenditure, and healthcare utilisation.

### 4.5. Time use for unpaid work and leisure activities

Times spent on all three measures of unpaid work were found to increase after retirement, with estimated coefficients of 0.47 - 0.78 hours per week. Although the effects of retirement on consumption were not observed, we found that satisfaction with leisure length enhanced (b: 3.58, SE: 1.33).

### 4.6. Lagged effects

To evaluate the longer-term or lagged effects of retirement on health and its mediating factors, we assessed retirement at years T-1 and T-2 on dependent variables at year T (Table 3 and 4). We found persistent increases, even one or two years after retirement, on exercise days, and time spent on volunteer and childcare, which can be habitual activities. Meanwhile, improvements in psychological health and psychological well-being were no longer observed when the two-year lagged effects of retirement were estimated. While we observed lagged protective effects of retirement on some of the physical health measures, this may be affected by changes in the probability of receiving health check-ups after retirement.

**Table 3.** Lagged effects of retirement on health and psychological well-being

| Dependent variable | Retirement at T-1 |  |  |  |  | Retirement at T-2 |  |  |  |  |
| --- | --- | --- | --- | --- | --- | --- | --- | --- | --- | --- |
|  | beta (SE) | Cragg-Donald<br>Wald F | Hansen J<br>(p-value) | Observations | N of id | beta (SE) | Cragg-Donald<br>Wald F | Hansen J<br>(p-value) | Observations | N of id |
| SRH | 0.03 (0.13) | 38.99 | 0.23 | 9,276 | 1,327 | -0.10* (0.05) | 24.29 | 0.37 | 8,198 | 1,239 |
| Psychosomatic symptoms | -0.15 (0.09) | 29.96 | 0.20 | 8,223 | 1,287 | -0.03 (0.02) | 18.30 | 0.06 | 7,158 | 1,177 |
| Somatic symptoms | -0.12 (0.09) | 30.09 | 0.51 | 8,218 | 1,287 | 0.02 (0.11) | 18.30 | 0.13 | 7,158 | 1,177 |
| Psychological distress | -0.19 (0.11) | 30.03 | 0.04 | 8,207 | 1,287 | 0.03 (0.14) | 18.32 | 0.03 | 7,143 | 1,176 |
| Life satisfaction | 0.74 (0.85) | 25.99 | 0.96 | 5,562 | 1,039 | 0.29 (0.23) | 16.68 | 0.60 | 5,122 | 1,000 |
| Blood pressure | -0.26 (0.37) | 13.87 | 0.47 | 5,224 | 978 | -0.18 (0.11) | 11.93 | 0.05 | 4,997 | 947 |
| Bone density | -0.06 (0.07) | 13.87 | 0.34 | 5,224 | 978 | -0.02 (0.02) | 11.93 | 0.22 | 4,997 | 947 |
| Heart | -0.79* (0.34) | 13.87 | 0.50 | 5,224 | 978 | -0.22* (0.09) | 11.93 | 0.20 | 4,997 | 947 |
| Anaemia | -0.12 (0.16) | 13.87 | 0.35 | 5,224 | 978 | -0.12* (0.06) | 11.93 | 0.46 | 4,997 | 947 |
| Liver | -0.31 (0.30) | 13.87 | 0.97 | 5,224 | 978 | -0.07 (0.07) | 11.93 | 0.36 | 4,997 | 947 |
| Kidney | -0.24 (0.21) | 13.87 | 0.35 | 5,224 | 978 | -0.10 (0.05) | 11.93 | 0.24 | 4,997 | 947 |
| Diabetes | 0.02 (0.27) | 13.87 | 0.22 | 5,224 | 978 | -0.13 (0.08) | 11.93 | 0.31 | 4,997 | 947 |
| Metabolism | 0.16 (0.22) | 13.87 | 0.11 | 5,224 | 978 | -0.00 (0.05) | 11.93 | 0.26 | 4,997 | 947 |
| Obesity | -0.06 (0.04) | 33.69 | 0.10 | 5,562 | 939 | -0.01 (0.01) | 22.62 | 0.08 | 4,770 | 744 |
| Overweight | -0.06 (0.12) | 33.69 | 0.45 | 5,562 | 939 | -0.01 (0.03) | 22.62 | 0.07 | 4,770 | 744 |

Note: Estimated coefficients and robust standard errors (SE) in parentheses; All models include the same controls as the main analysis; Full results are available upon request.

**Table 4.** Lagged effects of retirement on health behaviours, time use, and leisure activities

| Dependent variable | Retirement at T-1 |  |  |  |  | Retirement at T-2 |  |  |  |  |
| --- | --- | --- | --- | --- | --- | --- | --- | --- | --- | --- |
|  | beta (SE) | Cragg-Donald<br>Wald F | Hansen J<br>(p-value) | Observations | N of id | beta (SE) | Cragg-Donald<br>Wald F | Hansen J<br>(p-value) | Observations | N of id |
| BMI | -0.00 (0.02) | 33.69 | 0.93 | 5,562 | 939 | 0.00 (0.01) | 22.62 | 0.49 | 4,770 | 744 |
| Smoking | -0.24* (0.11) | 38.48 | 0.10 | 9,250 | 1,326 | -0.05 (0.04) | 24.36 | 0.24 | 8,174 | 1,238 |
| Alcohol | -0.17 (0.13) | 35.04 | 0.96 | 8,884 | 1,317 | -0.04 (0.04) | 21.56 | 0.57 | 7,802 | 1,205 |
| Sleep | 0.46 (0.33) | 33.49 | 0.01 | 8,571 | 1,300 | -0.03 (0.11) | 21.36 | 0.07 | 7,524 | 1,188 |
| Exercise | 1.75** (0.49) | 27.07 | 0.60 | 7,744 | 1,228 | 0.42** (0.15) | 19.91 | 0.31 | 7,060 | 1,146 |
| Health check-up | -0.45* (0.21) | 29.25 | 0.21 | 7,767 | 1,233 | 0.03 (0.07) | 19.18 | 0.84 | 7,069 | 1,148 |
| Healthcare use | 0.29 (0.21) | 29.58 | 0.83 | 7,760 | 1,232 | 0.07 (0.06) | 19.04 | 0.99 | 7,066 | 1,148 |
| Total health expenditure | 0.04 (0.03) | 28.91 | 0.92 | 8,595 | 1,286 | 0.01 (0.01) | 20.57 | 0.59 | 7,609 | 1,199 |
| Time: Volunteer | 0.74** (0.20) | 34.45 | 0.49 | 8,670 | 1,299 | 0.18* (0.07) | 19.90 | 0.39 | 7,692 | 1,214 |
| Time: Domestic work | 0.59* (0.28) | 36.72 | 0.21 | 8,861 | 1,307 | 0.08 (0.09) | 24.39 | 0.35 | 7,865 | 1,227 |
| Time: Childcare | 0.54* (0.23) | 15.46 | 0.85 | 6,034 | 1,020 | 0.10 (0.08) | 11.17 | 0.87 | 5,354 | 964 |
| Consumption: Eat-out | 0.00 (0.02) | 37.22 | 0.62 | 8,588 | 1,279 | -0.00 (0.01) | 23.78 | 0.31 | 7,679 | 1,204 |
| Consumption: Entertainment | 0.02 (0.02) | 31.91 | 0.90 | 8,384 | 1,269 | 0.01 (0.01) | 24.18 | 0.55 | 7,437 | 1,192 |
| Consumption: Social | -0.05 (0.04) | 28.60 | 0.52 | 8,621 | 1,284 | 0.00 (0.01) | 20.69 | 0.21 | 7,646 | 1,203 |
| Satisfaction: Leisure length | 2.64* (1.16) | 26.23 | 0.34 | 5,559 | 1,039 | 0.93** (0.34) | 15.68 | 0.43 | 5,119 | 999 |
| Satisfaction: Leisure activity | 0.39 (0.96) | 26.50 | 0.39 | 5,559 | 1,039 | 0.54 (0.29) | 16.13 | 0.28 | 5,119 | 1,000 |

Note: Estimated coefficients and robust standard errors (SE) in parentheses; All models include the same controls as the main analysis; Full results are available upon request.

### 4.7. Heterogeneity

To better understand the heterogeneous effects of retirement among different subgroups, some of the previous studies described earlier detected heterogeneity in retirement effects on health by subgroups, such as gender, education, occupation, and income, because health deterioration, health investment patterns, and retirement adjustment may vary among dissimilar individuals with different socioeconomic backgrounds. Based on the Grossman model (Grossman, 2000), there are rationales to believe in heterogeneity across socioeconomic groups, such as education, occupation, and income: The health gradient arises because the better-educated individuals produce health more efficiently, workers whose job requires intensive physical or mental burdens can undergo larger health deterioration, and the wealthier have more resources to invest in their health. Accordingly, we conducted additional analyses by income, occupation, and education to assess potential heterogeneity across these subgroups.

First, for income, we classified individuals into high- and low-income groups (i.e. higher or lower than the median of the sample), based on the average net personal pre-retirement income within the study period adjusted for prices. Remarkably, improvements in psychological health and subjective well-being, as well as time dedicated to unpaid work and consumption for health, were mostly found in the high-income group (Table 5 and 6). Although post-retirement increases in sleeping hours and exercise days were observed in the low-income group, adverse effects on smoking were observed among them, which we found the opposite in the high-income group. Second, for occupation, we assessed the heterogeneity of the effect of retirement from physically demanding jobs and non-physically demanding jobs, with physically demanding jobs defined as agriculture, forestry, fishery, mining, manufacturing, and protective service workers. The effects of retirement observed in our main analysis were mostly found among retirees of non-physically demanding jobs (Appendix Table A-6 and A-7). Finally, we assessed heterogeneity across education, classifying individuals into two groups of university graduates or higher and those with education lower than university graduates. We found some of the items in time use for unpaid work in both groups, but differences in other domains were less clear (Appendix Table A-8 and A-9).

**Table 5.**
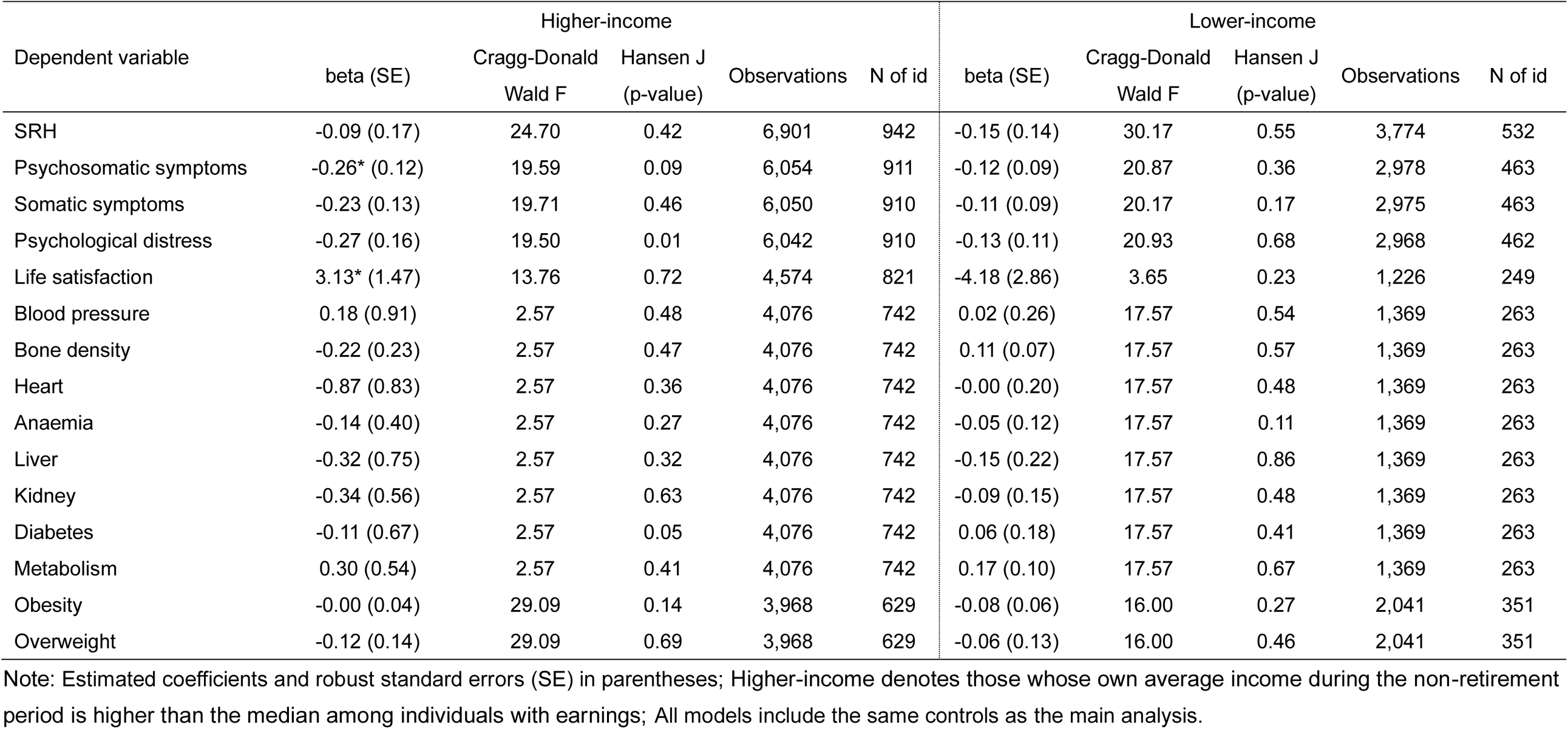
Heterogeneous effects of retirement by income on health and psychological well-being

**Table 6.** Heterogeneous effects of retirement by income on health behaviours, time use, and leisure activities

| Dependent variable | Higher-income |  |  |  |  | Lower-income |  |  |  |  |
| --- | --- | --- | --- | --- | --- | --- | --- | --- | --- | --- |
|  | beta (SE) | Cragg-Donald<br>Wald F | Hansen J<br>(p-value) | Observations | N of id | beta (SE) | Cragg-Donald<br>Wald F | Hansen J<br>(p-value) | Observations | N of id |
| BMI | -0.01 (0.03) | 29.09 | 0.93 | 3,968 | 629 | 0.06* (0.03) | 16.00 | 0.43 | 2,041 | 351 |
| Smoking | -0.86** (0.22) | 24.72 | 0.13 | 6,884 | 940 | 0.42** (0.13) | 30.28 | 0.46 | 3,762 | 532 |
| Alcohol | -0.17 (0.18) | 22.13 | 0.29 | 6,605 | 939 | -0.11 (0.14) | 23.65 | 0.54 | 3,130 | 465 |
| Sleep | 0.15 (0.43) | 21.60 | 0.43 | 6,059 | 896 | 1.16** (0.43) | 20.68 | 0.15 | 2,926 | 449 |
| Exercise | 1.38* (0.67) | 13.83 | 0.13 | 5,199 | 829 | 0.98** (0.36) | 25.13 | 0.63 | 2,931 | 443 |
| Health check-up | -0.59* (0.30) | 15.91 | 0.93 | 5,221 | 832 | -0.32 (0.20) | 25.06 | 0.90 | 2,928 | 444 |
| Healthcare use | 0.61 (0.34) | 15.81 | 0.43 | 5,208 | 832 | 0.06 (0.20) | 26.04 | 0.44 | 2,937 | 444 |
| Total health expenditure | 0.09* (0.04) | 20.32 | 0.68 | 6,459 | 925 | -0.01 (0.02) | 25.44 | 0.89 | 3,360 | 492 |
| Time: Volunteer | 1.11** (0.30) | 22.25 | 0.37 | 6,510 | 929 | 0.29 (0.15) | 24.20 | 0.49 | 3,437 | 501 |
| Time: Domestic work | 1.05** (0.41) | 25.49 | 0.64 | 6,208 | 902 | 0.45 (0.31) | 23.28 | 0.07 | 3,191 | 461 |
| Time: Childcare | 0.50* (0.22) | 18.29 | 0.45 | 4,800 | 774 | 0.44* (0.21) | 10.99 | 0.09 | 1,852 | 319 |
| Consumption: Eat-out | 0.00 (0.03) | 20.09 | 0.27 | 6,479 | 923 | 0.02 (0.02) | 24.65 | 0.32 | 3,303 | 483 |
| Consumption: Entertainment | 0.04 (0.03) | 20.56 | 0.03 | 6,247 | 910 | 0.03 (0.02) | 26.93 | 0.61 | 3,319 | 483 |
| Consumption: Social | -0.01 (0.06) | 17.22 | 0.75 | 6,491 | 922 | -0.01 (0.04) | 23.93 | 0.30 | 3,343 | 488 |
| Satisfaction: Leisure length | 4.20* (1.72) | 13.89 | 0.51 | 4,572 | 821 | -2.06 (3.53) | 3.63 | 0.08 | 1,222 | 249 |
| Satisfaction: Leisure activity | 1.97 (1.41) | 13.82 | 0.98 | 4,569 | 820 | 0.41 (2.75) | 3.70 | 0.93 | 1,224 | 249 |

Note: Estimated coefficients and robust standard errors (SE) in parentheses; Higher-income denotes those whose own average income during the non-retirement period is higher than the median among individuals with earnings; All models include the same controls as the main analysis.

## 5. Discussion

In this study, we aimed to evaluate the effects of retirement on health and potential mediators of the linkage between retirement and health. Based on our analysis using the instrumental variable fixed-effects model, we obtained three main findings. First, we observed improvements after retirement in psychological distress and subjective well-being, as well as increases in exercise habits and time spent on unpaid work. In contrast, the effects on health, measured by self-rated health and body mass index, were not observed. Second, the psychological benefits of retirement were no longer observed for longer durations after retirement. In contrast, healthy habits such as exercise and unpaid activities, such as volunteer work, continue. Third, the heterogeneous effects of retirement, particularly by income, were observed, suggesting that the improvements after retirement in the variables of interest occurred mostly among the higher-income group.

In terms of the psychological benefits of retirement, our findings are partly in line with many previous findings (Atalay & Barrett, 2014; Eibich, 2015; Gorry et al., 2018; Kolodziej & Garcia-Gomez, 2019; Nishimura et al., 2018; Rose, 2020). In addition, our findings on unpaid activities and increased exercise or physical activity are consistent with the existing studies (Eibich, 2015; Kampfen & Maurer, 2016; Kesavayuth et al., 2018).

Considering that psychological distress declined and satisfaction with leisure length and overall life improved, retirement worked as relief from a stressful working life (Bossé et al., 1991), rather than as a stressful life event (Minkler, 1981). One of the potential reasons for this is that working long hours is a serious issue in Japan, which can harm workers’ psychological health. In the worst case, long hours of work can lead to *karoshi* or death owing to overworking. Therefore, retirement from a tough working life may give individuals a restful feeling as well as more time available to take care of their health.

Our findings were not consistent with a study from the same country, which analysed the data of older birth cohorts than this study (Kajitani et al., 2016; Okamoto et al., 2018). Whilst this divergence may stem from methodological differences, there are possibly heterogeneities in work ethics and style across generations. Compared with the earlier generations, Japanese men born around World War II, who were the majority of the sample of this study, tended to spend a work-oriented life with high work centrality with increasing employed labourers and undergoing a period of high economic growth (Japan Institute for Labour Policy and Training, 2007), which can impose heavy workloads during their career life; thus, retirement may be viewed as a release from a stressful working life. Therefore, even by studies within the same country, there is no wonder if the results are mixed when the data derived from different generations are analysed. Although this requires further investigation, exploring potential heterogeneity by work ethics and work styles may be useful to understand why findings from existing studies are controversial. Therefore, meta-analysis or cross-country studies are required, covering regions and cohorts in various settings, to detect the institutional or cultural reasons behind the heterogeneous health effects of retirement observed.

Even though the positive psychological effects of retirement were observed, the effects were no longer significant in longer periods after retirement. This could be because retired individuals may enjoy a ‘honeymoon period’ following retirement, where individuals were engaged in activities that they had to forgo owing to work-related constraints, or a brief respite after a long period of time as a labour force participant. These psychological benefits may be temporal; after a while, they may start to adjust their retirement life and return to a pre-retirement level. Interestingly, in contrast, retirees continued to engage in healthy habits such as exercise and unpaid activities such as volunteer work even for a longer duration after retirement, which may lead to an eventual decline in the likelihood of being obese. This may suggest that a habit, once formed, remains held in the post-retirement life, and improvements in physical health can be consequently observed in the long run.

However, among the measures for healthcare utilisation, we found that the probabilities of receiving health check-ups declined after retirement. One of the most important reasons for this is that access to annual health check-ups changes after retirement. Employees are able to receive health check-ups in their workplace, as employers are required by law to provide annual health check-ups for them (Okamura et al., 2014). Contrary, many of the retirees receive health check-ups in their municipality of residence, some of which impose out-of-pocket expenses on individuals. Therefore, long-term health effects may be of concern due to forgone health check-ups by retirees, delaying disease detection in its earliest stages.

Contrary to our findings, one previous study reported that they found longer-term effects of retirement on mental health, but no short-term effects (Heller-Sahlgren, 2017). It is not entirely clear why different results were observed; this may be driven by any institutional or cultural differences between regions (e.g. how they worked until retirement or lived their retirement lives) as well as methodological differences (e.g. measures of mental health), which require further investigation.

Those with higher pre-retirement income benefited from retirement compared to the lower-income group. For this, there were at least two possible interpretations. One thing is that the higher-income group had more financial resources to invest in for their health, as the level of employees’ pension benefits is associated with pre-retirement income, which also enables them to adjust to their retirement life well (Wang, 2013). Another explanation, being related to a continuous theory (Wang, 2013), is that those with higher non-cognitive skills, which are reported to be linked to higher earnings (Heckman et al., 2006), may better manage to find new roles after retirement through unpaid activities, and live their successful retirement lives. Moreover, non-cognitive skills have been reported to be positively associated with good health behaviours (Chiteji, 2010), which enforces the first interpretation that the higher-income group was endowed with non-financial resources.

From our findings, there are three possible policy implications for better living conditions in older populations before and after retirement. First, it is essential to enhance work environments to avoid excessive health deterioration. Even if health improvements are observed after retirement, the psychological benefits may vanish in a short period of time, and severe health losses owing to hard work cannot be recovered. Policies for attaining this would include protecting workers from undesired long working hours and sustaining a good work-life balance.

Second, as a habit is persistent after retirement, it is important to assist individuals in formalising good habits before and after retirement. Also, it is essential to enable retirees to easily access health check-ups by reducing associated costs (e.g. out-of-pocket expenses, transportation, and appointment). Moreover, creating opportunities for individuals to find new roles in their post-retirement lives is also important. For this, in addition to a work-life balance enabling an engagement in those activities even before retirement, lessons learned from promoting activities for social engagement of middle- and old-age populations should be effectively shared among stakeholders to encourage retirees to stay active.

Third, social policies to mitigate socioeconomic disparities in health are important, enabling even low-income individuals to enjoy a healthy retirement life. As socioeconomic and health disparities are being generated, not only during a specific moment of life, but also throughout life, financial and non-financial (e.g. education and in-kind benefits) support from the cradle to the grave for those living under adverse socioeconomic circumstances are necessary.

Finally, this study has several limitations which should be considered when interpreting our findings. First, while we presented estimated lagged effects of retirement on dependent variables, we were unable to assess effects for much longer durations (e.g. 10 years or more). Despite its importance, it may not be easy to estimate ultralong-run effects since there would be many other ‘noises’ that affect one’s health status. Second, we were unable to provide estimates for specific subgroups (e.g. women and self-employed individuals) because of their dissimilar retirement behaviours to male employees, which makes identification by our IV approach difficult. Therefore, it is necessary to assess heterogeneity across a wide variety of subgroups to better understand the mechanisms behind a retirement-and-health relationship. Third, although we covered various types of outcome variables, the JHPS/KHPS included only limited health measures, and by analysing diverse health outcomes, we might have observed their changes associated with retirement, which enriches interpretations of the results. Fourth, as retirement is a complex procedure, its alternative definitions (e.g. partial retirement) may provide different results; however, we were unable to test this due to the data restriction.

In conclusion, this study investigates the effects of retirement on health and potential mediators of the linkage between retirement and health using an instrumental variable fixed-effects approach, suggesting that retirement improves psychological distress and subjective well-being as well as increases exercise habits and time spent on unpaid work. Together with results from lagged and subgroup analyses, policymakers should consider the mechanisms behind retirement-and-health relationships along with the potential heterogeneous effects when increasing retirement ages, incorporating additional policies necessary for healthy retirement lives.

## Data Availability

The data that support the findings of this study are available from the Panel Data Research Center of Keio University. Data are available at https://www.pdrc.keio.ac.jp/en/paneldata/datasets/jhpskhps/ with the permission of the Panel Data Research Center of Keio University.

## Acknowledgements

We thank the Panel Data Research Center of Keio University for providing the data, the Japan Household Panel Survey, for a secondary analysis in this study. We are also grateful for helpful feedbacks from Dr Rei Goto, Dr Hirotaka Kato, Mr Shingo Kasahara, Mr Tatsunari Miyayama, and Ms Tomomi Maeda at Keio University.

## Supplementary material

**Appendix Table A-1.**
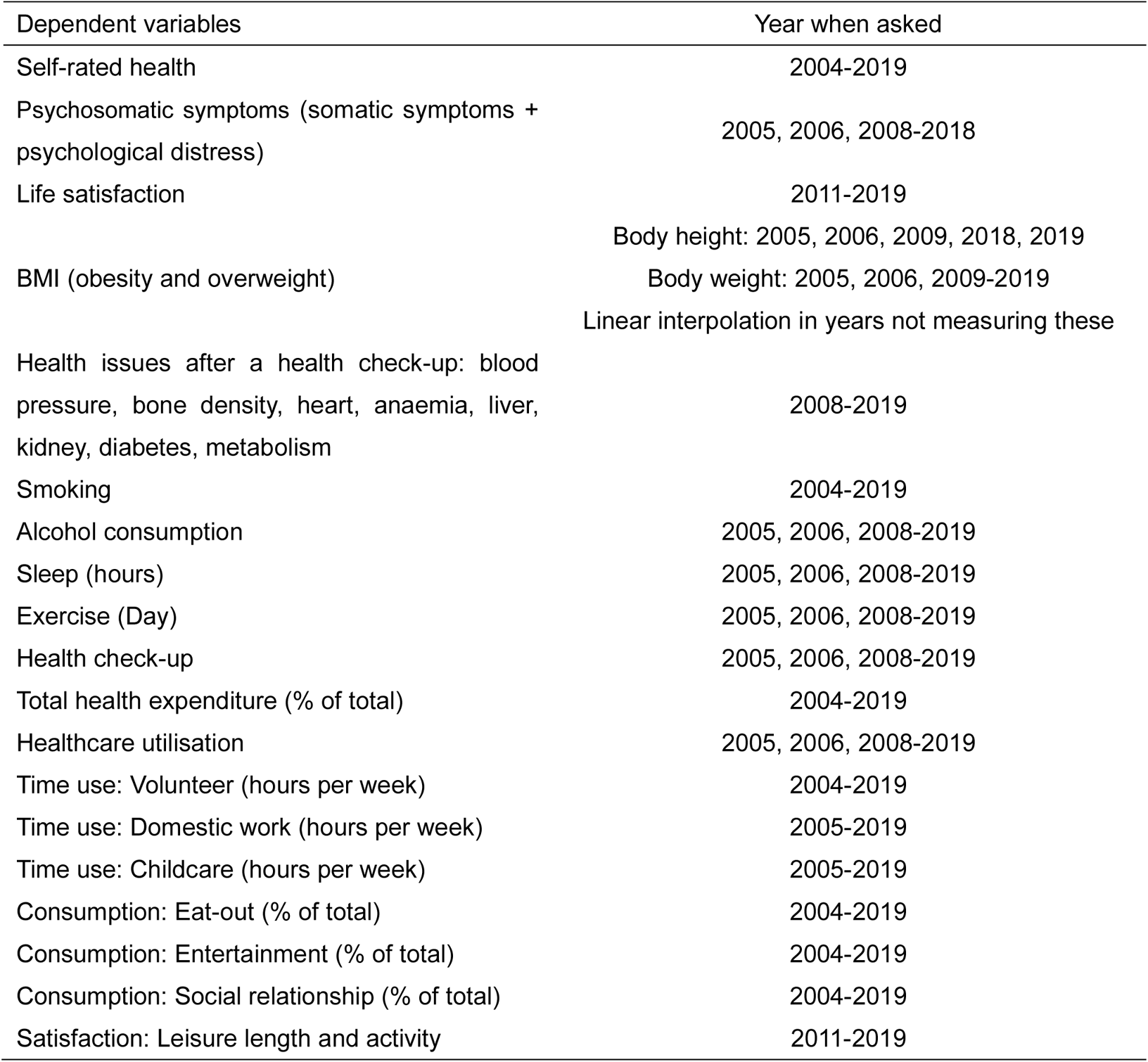
Years when dependent variables are measured

| Dependent variables | Year when asked |
| --- | --- |
| Self-rated health | 2004-2019 |
| Psychosomatic symptoms (somatic symptoms + psychological distress) | 2005, 2006, 2008-2018 |
| Life satisfaction | 2011-2019 |
|  | Body height: 2005, 2006, 2009, 2018, 2019 |
| BMI (obesity and overweight) | Body weight: 2005, 2006, 2009-2019 |
|  | Linear interpolation in years not measuring these |
| Health issues after a health check-up: blood pressure, bone density, heart, anaemia, liver, kidney, diabetes, metabolism | 2008-2019 |
| Smoking | 2004-2019 |
| Alcohol consumption | 2005, 2006, 2008-2019 |
| Sleep (hours) | 2005, 2006, 2008-2019 |
| Exercise (Day) | 2005, 2006, 2008-2019 |
| Health check-up | 2005, 2006, 2008-2019 |
| Total health expenditure (% of total) | 2004-2019 |
| Healthcare utilisation | 2005, 2006, 2008-2019 |
| Time use: Volunteer (hours per week) | 2004-2019 |
| Time use: Domestic work (hours per week) | 2005-2019 |
| Time use: Childcare (hours per week) | 2005-2019 |
| Consumption: Eat-out (% of total) | 2004-2019 |
| Consumption: Entertainment (% of total) | 2004-2019 |
| Consumption: Social relationship (% of total) | 2004-2019 |
| Satisfaction: Leisure length and activity | 2011-2019 |

**Appendix Table A-2.** Full results of IV estimations for subjective health and psychological well-being

|  | SRH | Psychosomatic symptoms | Somatic symptoms | Psychological distress | Life satisfaction |
| --- | --- | --- | --- | --- | --- |
| Retire | -0.07<br>(0.10) | -0.19**<br>(0.07) | -0.17*<br>(0.07) | -0.23**<br>(0.09) | 2.17*<br>(1.06) |
| Age | -0.02<br>(0.02) | -0.06**<br>(0.02) | -0.05**<br>(0.02) | -0.09**<br>(0.02) | 0.61**<br>(0.19) |
| Age<sup>2</sup> | 0.03<br>(0.02) | 0.06**<br>(0.01) | 0.05**<br>(0.01) | 0.09**<br>(0.02) | -0.45**<br>(0.15) |
| Single | -0.03<br>(0.04) | 0.03<br>(0.02) | 0.04<br>(0.03) | 0.01<br>(0.03) | -0.20<br>(0.30) |
| Living alone | 0.08**<br>(0.03) | -0.04*<br>(0.02) | -0.03<br>(0.02) | -0.05*<br>(0.02) | -0.21<br>(0.20) |
| Home ownership | -0.07<br>(0.04) | 0.03<br>(0.02) | 0.00<br>(0.02) | 0.07*<br>(0.03) | -0.05<br>(0.32) |
| Individual FE | Yes | Yes | Yes | Yes | Yes |
| Year FE | Yes | Yes | Yes | Yes | Yes |
| Prefecture-by-scale FE | Yes | Yes | Yes | Yes | Yes |
| Cragg-Donald Wald F statistic: | 61.34 | 47.33 | 46.42 | 47.45 | 20.46 |
| Hansen J statistic (p-value) | 0.668 | 0.0628 | 0.199 | 0.0525 | 0.428 |
| Observations | 10,682 | 9,037 | 9,030 | 9,015 | 5,801 |
| Number of id | 1,469 | 1,370 | 1,369 | 1,368 | 1,068 |

Note: Estimated coefficients and robust standard errors in parentheses; Prefecture-by-scale dummy is partialled out from all the other variables, using the Stata's *partial* option; Singleton groups are not used for estimations; FE denotes fixed-effects.

**Appendix Table A-3.** Full results of IV estimations for physical health

|  | Blood<br>pressure | Bone<br>density | Heart | Anaemia | Liver | Kidney | Diabetes | Metabolomes | Obesity | Overweight |
| --- | --- | --- | --- | --- | --- | --- | --- | --- | --- | --- |
| Retire | 0.12<br>(0.27) | -0.00<br>(0.05) | -0.37<br>(0.20) | -0.06<br>(0.12) | -0.28<br>(0.22) | -0.19<br>(0.16) | -0.10<br>(0.20) | 0.18<br>(0.16) | -0.05<br>(0.03) | -0.10<br>(0.09) |
| Age | 0.03<br>(0.05) | -0.01<br>(0.01) | -0.08*<br>(0.04) | 0.01<br>(0.02) | -0.07<br>(0.04) | -0.05<br>(0.03) | -0.03<br>(0.04) | 0.05<br>(0.03) | 0.00<br>(0.01) | -0.01<br>(0.02) |
| Age<sup>2</sup> | -0.01<br>(0.04) | 0.01<br>(0.01) | 0.06<br>(0.03) | 0.00<br>(0.02) | 0.06<br>(0.03) | 0.05*<br>(0.02) | 0.04<br>(0.03) | -0.03<br>(0.02) | -0.00<br>(0.00) | 0.00<br>(0.01) |
| Single | -0.02<br>(0.05) | -0.01<br>(0.01) | 0.00<br>(0.04) | -0.03<br>(0.02) | -0.05<br>(0.06) | -0.02<br>(0.04) | -0.01<br>(0.05) | -0.05<br>(0.04) | 0.00<br>(0.01) | -0.04<br>(0.04) |
| Living alone | -0.09<br>(0.05) | 0.02<br>(0.02) | 0.04<br>(0.04) | 0.04<br>(0.02) | 0.02<br>(0.05) | 0.02<br>(0.03) | 0.02<br>(0.04) | -0.03<br>(0.03) | -0.01<br>(0.01) | -0.01<br>(0.03) |
| Home ownership | -0.14*<br>(0.07) | 0.03<br>(0.03) | -0.02<br>(0.04) | -0.00<br>(0.01) | -0.03<br>(0.06) | 0.02<br>(0.05) | 0.01<br>(0.05) | -0.04<br>(0.03) | -0.01<br>(0.02) | -0.04<br>(0.04) |
| Individual FE | Yes | Yes | Yes | Yes | Yes | Yes | Yes | Yes | Yes | Yes |
| Year FE | Yes | Yes | Yes | Yes | Yes | Yes | Yes | Yes | Yes | Yes |
| Prefecture-by-scale<br>FE | Yes | Yes | Yes | Yes | Yes | Yes | Yes | Yes | Yes | Yes |
| Cragg-Donald Wald F<br>statistic: | 22.45 | 22.45 | 22.45 | 22.45 | 22.45 | 22.45 | 22.45 | 22.45 | 54.26 | 54.26 |
| Hansen J statistic (p-value) | 0.373 | 0.546 | 0.420 | 0.0740 | 0.430 | 0.615 | 0.0271 | 0.321 | 0.396 | 0.875 |
| Observations | 5,446 | 5,446 | 5,446 | 5,446 | 5,446 | 5,446 | 5,446 | 5,446 | 6,012 | 6,012 |
| Number of id | 1,004 | 1,004 | 1,004 | 1,004 | 1,004 | 1,004 | 1,004 | 1,004 | 975 | 975 |

Note: Estimated coefficients and robust standard errors in parentheses; Prefecture-by-scale dummy is partialled out from all the other variables, using the Stata's *partial* option; Singleton groups are not used for estimations; FE denotes fixed-effects.

**Appendix Table A-4.** Full results of IV estimations for subjective well-being and time use for unpaid works

|  | BMI | Smoking | Alcohol | Sleep | Exercise | Check-up | Healthcare<br>use | Total health<br>expenditure |
| --- | --- | --- | --- | --- | --- | --- | --- | --- |
| Retire | 0.01<br>(0.02) | -0.19*<br>(0.09) | -0.13<br>(0.10) | 0.41<br>(0.27) | 1.09**<br>(0.31) | -0.33*<br>(0.15) | 0.24<br>(0.16) | 0.03<br>(0.02) |
| Age | 0.01<br>(0.00) | -0.05**<br>(0.02) | -0.01<br>(0.02) | 0.06<br>(0.06) | 0.19**<br>(0.07) | 0.01<br>(0.03) | 0.08*<br>(0.03) | 0.01*<br>(0.00) |
| Age<sup>2</sup> | -0.01**<br>(0.00) | 0.03*<br>(0.02) | -0.01<br>(0.02) | -0.05<br>(0.04) | -0.15**<br>(0.05) | 0.01<br>(0.02) | -0.06*<br>(0.02) | -0.01<br>(0.00) |
| Single | -0.00<br>(0.01) | 0.05<br>(0.04) | 0.01<br>(0.03) | 0.03<br>(0.10) | -0.09<br>(0.09) | 0.09<br>(0.05) | -0.01<br>(0.06) | -0.01<br>(0.01) |
| Living alone | -0.01<br>(0.01) | -0.01<br>(0.02) | 0.04<br>(0.03) | -0.15*<br>(0.07) | 0.04<br>(0.07) | -0.04<br>(0.03) | -0.02<br>(0.04) | -0.01<br>(0.00) |
| Home ownership | -0.01<br>(0.01) | 0.02<br>(0.03) | 0.04<br>(0.03) | 0.10<br>(0.09) | 0.16<br>(0.10) | -0.08*<br>(0.04) | -0.12**<br>(0.04) | -0.00<br>(0.01) |
| Individual FE | Yes | Yes | Yes | Yes | Yes | Yes | Yes | Yes |
| Year FE | Yes | Yes | Yes | Yes | Yes | Yes | Yes | Yes |
| Prefecture-by-scale FE | Yes | Yes | Yes | Yes | Yes | Yes | Yes | Yes |
| Cragg-Donald Wald F<br>statistic: | 54.26 | 61.60 | 55.10 | 52.08 | 46 | 48.63 | 49.46 | 53.90 |
| Hansen J statistic (p-value) | 0.591 | 0.519 | 0.589 | 0.128 | 0.207 | 0.951 | 0.284 | 0.786 |
| Observations | 6,012 | 10,653 | 9,739 | 8,986 | 8,130 | 8,149 | 8,145 | 9,825 |
| Number of id | 975 | 1,467 | 1,399 | 1,341 | 1,269 | 1,273 | 1,273 | 1,412 |

Note: Estimated coefficients and robust standard errors in parentheses; Prefecture-by-scale dummy is partialled out from all the other variables, using the Stata's *partial* option; Singleton groups are not used for estimations; FE denotes fixed-effects.

**Appendix Table A-5.** Full results of IV estimations for time use and leisure activities

|  | Time use |  |  | Consumption |  |  | Satisfaction |  |
| --- | --- | --- | --- | --- | --- | --- | --- | --- |
|  | Volunteer | Domestic work | Childcare | Eat-out | Entertainment | Social | Leisure length | Leisure activities |
| Retire | 0.60**<br>(0.13) | 0.78**<br>(0.23) | 0.47**<br>(0.15) | 0.01<br>(0.01) | 0.02<br>(0.02) | -0.01<br>(0.03) | 3.58**<br>(1.33) | 1.66<br>(1.11) |
| Age | 0.09**<br>(0.03) | 0.11*<br>(0.05) | 0.05<br>(0.03) | 0.00<br>(0.00) | 0.01<br>(0.00) | 0.01<br>(0.01) | 0.85**<br>(0.24) | 0.48*<br>(0.20) |
| Age<sup>2</sup> | -0.09**<br>(0.02) | -0.08*<br>(0.04) | -0.05*<br>(0.03) | -0.00<br>(0.00) | -0.00<br>(0.00) | -0.01<br>(0.01) | -0.66**<br>(0.19) | -0.36*<br>(0.16) |
| Single | 0.00<br>(0.03) | 0.27**<br>(0.08) | -0.00<br>(0.03) | 0.00<br>(0.01) | 0.01<br>(0.01) | -0.01<br>(0.01) | 0.76*<br>(0.32) | 0.34<br>(0.30) |
| Living alone | -0.04<br>(0.02) | 0.23**<br>(0.06) | -0.02<br>(0.02) | 0.01<br>(0.00) | -0.01<br>(0.00) | 0.01<br>(0.01) | -0.96**<br>(0.30) | -0.58*<br>(0.27) |
| Home ownership | -0.03<br>(0.04) | 0.02<br>(0.08) | -0.10*<br>(0.05) | 0.00<br>(0.00) | -0.00<br>(0.01) | 0.01<br>(0.01) | -0.45<br>(0.44) | 0.10<br>(0.41) |
| Individual FE | Yes | Yes | Yes | Yes | Yes | Yes | Yes | Yes |
| Year FE | Yes | Yes | Yes | Yes | Yes | Yes | Yes | Yes |
| Prefecture-by-scale FE | Yes | Yes | Yes | Yes | Yes | Yes | Yes | Yes |
| Cragg-Donald Wald F statistic: | 54.59 | 59.20 | 34.37 | 55.05 | 58.38 | 49.09 | 20.55 | 20.61 |
| Hansen J statistic (p-value) | 0.279 | 0.144 | 0.898 | 0.257 | 0.0163 | 0.82 | 0.103 | 0.791 |
| Observations | 9,955 | 9,400 | 6,656 | 9,788 | 9,571 | 9,839 | 5,795 | 5,794 |
| Number of id | 1,426 | 1,359 | 1,091 | 1,401 | 1,388 | 1,405 | 1,068 | 1,067 |

Note: Estimated coefficients and robust standard errors in parentheses; Prefecture-by-scale dummy is partialled out from all the other variables, using the Stata's *partial* option; Singleton groups are not used for estimations; FE denotes fixed-effects.

**Appendix Table A-6:** Heterogeneous effects of retirement by occupation on health and psychological well-being

| Dependent variable | Physical demanding jobs |  |  |  |  | Non-physical demanding jobs |  |  |  |  |
| --- | --- | --- | --- | --- | --- | --- | --- | --- | --- | --- |
|  | beta (SE) | Cragg-Donald<br>Wald F | Hansen J<br>(p-value) | Observations | N of id | beta (SE) | Cragg-Donald<br>Wald F | Hansen J<br>(p-value) | Observations | N of id |
| SRH | 0.06 (0.20) | 16.87 | 0.28 | 2,951 | 499 | -0.02 (0.12) | 42.19 | 0.92 | 7,530 | 1,141 |
| Psychosomatic symptoms | -0.50** (0.19) | 12.75 | 0.21 | 2,499 | 452 | -0.10 (0.08) | 32.41 | 0.04 | 6,377 | 1,067 |
| Somatic symptoms | -0.45* (0.19) | 12.41 | 0.28 | 2,496 | 451 | -0.08 (0.08) | 31.94 | 0.16 | 6,372 | 1,066 |
| Psychological distress | -0.62** (0.23) | 12.92 | 0.25 | 2,494 | 452 | -0.11 (0.10) | 32.42 | 0.01 | 6,362 | 1,065 |
| Life satisfaction | 0.46 (1.44) | 7.88 | 0.01 | 1,507 | 329 | 2.49* (1.24) | 14.81 | 0.48 | 4,178 | 803 |
| Blood pressure | -0.18 (0.79) | 3.46 | 0.78 | 1,427 | 292 | 0.01 (0.29) | 19.45 | 0.35 | 3,909 | 776 |
| Bone density | 0.17 (0.18) | 3.46 | 0.57 | 1,427 | 292 | 0.03 (0.04) | 19.45 | 0.10 | 3,909 | 776 |
| Heart | -0.19 (0.53) | 3.46 | 0.89 | 1,427 | 292 | -0.24 (0.21) | 19.45 | 0.08 | 3,909 | 776 |
| Anaemia | 0.27 (0.42) | 3.46 | 0.55 | 1,427 | 292 | -0.10 (0.13) | 19.45 | 0.03 | 3,909 | 776 |
| Liver | -0.28 (0.58) | 3.46 | 0.35 | 1,427 | 292 | -0.32 (0.24) | 19.45 | 0.65 | 3,909 | 776 |
| Kidney | 0.20 (0.31) | 3.46 | 0.91 | 1,427 | 292 | -0.21 (0.18) | 19.45 | 0.23 | 3,909 | 776 |
| Diabetes | -0.65 (0.71) | 3.46 | 0.77 | 1,427 | 292 | 0.01 (0.20) | 19.45 | 0.07 | 3,909 | 776 |
| Metabolism | 0.13 (0.42) | 3.46 | 0.61 | 1,427 | 292 | 0.09 (0.17) | 19.45 | 0.64 | 3,909 | 776 |
| Obesity | -0.08 (0.07) | 12.00 | 0.30 | 1,608 | 302 | -0.02 (0.04) | 37.75 | 0.76 | 4,286 | 743 |
| Overweight | 0.01 (0.16) | 12.00 | 0.72 | 1,608 | 302 | -0.09 (0.11) | 37.75 | 0.80 | 4,286 | 743 |

Note: Estimated coefficients and robust standard errors (SE) in parentheses; Physical demanding jobs include jobs in agriculture, forestry, fishery, mining, manufacturing, and protective service works; All models include the same controls as the main analysis; Full results are available upon request.

**Appendix Table A-7.** Heterogeneous effects of retirement by occupation on health behaviours, time use, and leisure activities

| Dependent variable | Physical demanding jobs |  |  |  |  | Non-physical demanding jobs |  |  |  |  |
| --- | --- | --- | --- | --- | --- | --- | --- | --- | --- | --- |
|  | beta (SE) | Cragg-Donald<br>Wald F | Hansen J<br>(p-value) | Observations | N of id | beta (SE) | Cragg-Donald<br>Wald F | Hansen J<br>(p-value) | Observations | N of id |
| BMI | 0.06 (0.04) | 12.00 | 0.59 | 1,608 | 302 | -0.02 (0.02) | 37.75 | 0.75 | 4,286 | 743 |
| Smoking | -0.09 (0.16) | 17.15 | 0.23 | 2,943 | 498 | -0.26* (0.12) | 42.04 | 0.32 | 7,509 | 1,140 |
| Alcohol | -0.11 (0.17) | 15.49 | 0.63 | 2,691 | 467 | -0.24 (0.13) | 37.64 | 0.50 | 6,884 | 1,089 |
| Sleep | 0.50 (0.60) | 11.10 | 0.71 | 2,452 | 439 | 0.51 (0.31) | 36.96 | 0.29 | 6,374 | 1,038 |
| Exercise | 0.23 (0.66) | 12.13 | 0.04 | 2,319 | 418 | 1.63** (0.40) | 32.49 | 0.49 | 5,686 | 977 |
| Health check-up | -0.38 (0.27) | 13.51 | 0.05 | 2,319 | 418 | -0.34 (0.18) | 33.60 | 0.17 | 5,700 | 979 |
| Healthcare use | 0.17 (0.32) | 13.63 | 0.99 | 2,320 | 419 | 0.15 (0.19) | 34.46 | 0.09 | 5,696 | 979 |
| Total health expenditure | 0.03 (0.04) | 13.36 | 0.32 | 2,674 | 466 | 0.03 (0.02) | 37.35 | 0.98 | 6,955 | 1,094 |
| Time: Volunteer | 0.29 (0.19) | 13.33 | 0.04 | 2,765 | 479 | 0.61** (0.17) | 38.25 | 0.01 | 7,004 | 1,109 |
| Time: Domestic work | -0.34 (0.45) | 13.56 | 0.12 | 2,625 | 448 | 0.82** (0.27) | 45.11 | 0.49 | 6,621 | 1,061 |
| Time: Childcare | 0.76* (0.34) | 7.70 | 1.00 | 1,820 | 339 | 0.37* (0.16) | 29.45 | 0.92 | 4,710 | 855 |
| Consumption: Eat-out | -0.00 (0.03) | 14.84 | 0.89 | 2,682 | 467 | 0.01 (0.02) | 38.82 | 0.26 | 6,926 | 1,089 |
| Consumption: Entertainment | 0.03 (0.04) | 12.60 | 0.92 | 2,593 | 458 | 0.04 (0.02) | 42.87 | 0.01 | 6,783 | 1,076 |
| Consumption: Social | -0.11 (0.08) | 10.17 | 0.38 | 2,675 | 467 | 0.03 (0.04) | 36.07 | 0.61 | 6,973 | 1,094 |
| Satisfaction: Leisure length | 1.38 (2.00) | 7.92 | 0.03 | 1,504 | 329 | 3.91* (1.58) | 14.76 | 0.85 | 4,175 | 803 |
| Satisfaction: Leisure activity | 1.12 (1.80) | 7.88 | 0.31 | 1,505 | 329 | 2.28 (1.37) | 14.89 | 0.83 | 4,173 | 802 |

Note: Estimated coefficients and robust standard errors (SE) in parentheses; Physical demanding jobs include jobs in agriculture, forestry, fishery, mining, manufacturing, and protective service works; All models include the same controls as the main analysis; Full results are available upon request.

**Appendix Table A-8:** Heterogeneous effects of retirement by education on health and psychological well-being

| Dependent variable | University or higher |  |  |  |  | Lower than university |  |  |  |  |
| --- | --- | --- | --- | --- | --- | --- | --- | --- | --- | --- |
|  | beta (SE) | Cragg-Donald<br>Wald F | Hansen J<br>(p-value) | Observations | N of id | beta (SE) | Cragg-Donald<br>Wald F | Hansen J<br>(p-value) | Observations | N of id |
| SRH | 0.04 (0.13) | 40.29 | 0.30 | 3,919 | 537 | -0.22 (0.17) | 24.29 | 0.09 | 6,484 | 891 |
| Psychosomatic symptoms | -0.03 (0.08) | 32.56 | 0.49 | 3,346 | 509 | -0.26* (0.11) | 18.01 | 0.00 | 5,456 | 821 |
| Somatic symptoms | -0.04 (0.09) | 31.57 | 0.51 | 3,343 | 508 | -0.20 (0.12) | 17.78 | 0.01 | 5,453 | 821 |
| Psychological distress | -0.03 (0.11) | 32.77 | 0.43 | 3,342 | 509 | -0.33* (0.15) | 18.05 | 0.00 | 5,438 | 819 |
| Life satisfaction | 1.80 (1.11) | 18.60 | 0.55 | 2,311 | 408 | 2.45 (2.01) | 5.94 | 0.33 | 3,319 | 625 |
| Blood pressure | 0.08 (0.24) | 25.27 | 0.31 | 2,103 | 395 | 0.16 (0.47) | 7.84 | 0.70 | 3,181 | 578 |
| Bone density | -0.01 (0.04) | 25.27 | 0.23 | 2,103 | 395 | 0.00 (0.09) | 7.84 | 0.91 | 3,181 | 578 |
| Heart | -0.25 (0.21) | 25.27 | 0.28 | 2,103 | 395 | -0.36 (0.31) | 7.84 | 0.85 | 3,181 | 578 |
| Anaemia | -0.03 (0.12) | 25.27 | 0.06 | 2,103 | 395 | -0.12 (0.20) | 7.84 | 0.48 | 3,181 | 578 |
| Liver | -0.27 (0.20) | 25.27 | 0.74 | 2,103 | 395 | -0.27 (0.39) | 7.84 | 0.46 | 3,181 | 578 |
| Kidney | -0.15 (0.15) | 25.27 | 0.77 | 2,103 | 395 | -0.14 (0.24) | 7.84 | 0.72 | 3,181 | 578 |
| Diabetes | 0.01 (0.18) | 25.27 | 0.36 | 2,103 | 395 | -0.71 (0.42) | 7.84 | 0.07 | 3,181 | 578 |
| Metabolism | 0.01 (0.17) | 25.27 | 0.78 | 2,103 | 395 | 0.15 (0.24) | 7.84 | 0.03 | 3,181 | 578 |
| Obesity | -0.02 (0.03) | 27.04 | 0.07 | 2,213 | 368 | -0.04 (0.05) | 27.87 | 0.95 | 3,645 | 577 |
| Overweight | -0.24 (0.14) | 27.04 | 0.51 | 2,213 | 368 | 0.01 (0.12) | 27.87 | 0.98 | 3,645 | 577 |

Note: Estimated coefficients and robust standard errors (SE) in parentheses; All models include the same controls as the main analysis; Full results are available upon request.

**Appendix Table A-9.** Heterogeneous effects of retirement by education on health behaviours, time use, and leisure activities

| Dependent variable | University or higher |  |  |  |  | Lower than university |  |  |  |  |
| --- | --- | --- | --- | --- | --- | --- | --- | --- | --- | --- |
|  | beta (SE) | Cragg-Donald<br>Wald F | Hansen J<br>(p-value) | Observations | N of id | beta (SE) | Cragg-Donald<br>Wald F | Hansen J<br>(p-value) | Observations | N of id |
| BMI | -0.02 (0.02) | 27.04 | 0.71 | 2,213 | 368 | 0.02 (0.02) | 27.87 | 0.60 | 3,645 | 577 |
| Smoking | -0.20 (0.13) | 40.09 | 0.88 | 3,910 | 535 | -0.17 (0.13) | 24.62 | 0.23 | 6,467 | 891 |
| Alcohol | 0.21 (0.13) | 36.74 | 0.72 | 3,614 | 520 | -0.28 (0.16) | 21.01 | 0.84 | 5,864 | 838 |
| Sleep | 0.78* (0.32) | 37.26 | 0.80 | 3,356 | 503 | 0.11 (0.44) | 19.01 | 0.14 | 5,396 | 800 |
| Exercise | 0.65 (0.37) | 35.00 | 0.79 | 2,935 | 476 | 1.79** (0.55) | 18.10 | 0.09 | 4,971 | 754 |
| Health check-up | -0.14 (0.18) | 35.95 | 0.53 | 2,945 | 479 | -0.52* (0.23) | 19.63 | 0.47 | 4,983 | 755 |
| Healthcare use | -0.04 (0.19) | 35.11 | 0.56 | 2,945 | 478 | 0.51 (0.26) | 20.59 | 0.58 | 4,977 | 756 |
| Total health expenditure | 0.02 (0.02) | 38.33 | 0.99 | 3,647 | 519 | 0.03 (0.03) | 19.94 | 0.74 | 5,931 | 854 |
| Time: Volunteer | 0.80** (0.20) | 37.77 | 0.29 | 3,703 | 531 | 0.46* (0.19) | 20.33 | 0.29 | 5,990 | 855 |
| Time: Domestic work | 0.68* (0.30) | 41.99 | 0.54 | 3,501 | 509 | 0.68 (0.35) | 21.34 | 0.15 | 5,645 | 809 |
| Time: Childcare | 0.28 (0.26) | 12.21 | 0.02 | 2,619 | 419 | 0.61** (0.21) | 17.56 | 0.16 | 3,847 | 640 |
| Consumption: Eat-out | -0.00 (0.02) | 41.25 | 0.44 | 3,638 | 515 | 0.01 (0.02) | 19.27 | 0.56 | 5,906 | 847 |
| Consumption: Entertainment | 0.02 (0.02) | 41.20 | 0.07 | 3,560 | 513 | 0.02 (0.02) | 21.62 | 0.11 | 5,772 | 836 |
| Consumption: Social | 0.05 (0.04) | 38.71 | 0.70 | 3,647 | 519 | -0.06 (0.05) | 16.02 | 1.00 | 5,941 | 846 |
| Satisfaction: Leisure length | 2.48 (1.28) | 18.58 | 0.31 | 2,311 | 408 | 4.73 (2.63) | 6.03 | 0.05 | 3,313 | 625 |
| Satisfaction: Leisure activity | 0.72 (1.10) | 18.63 | 0.65 | 2,311 | 407 | 3.04 (2.17) | 6.03 | 0.19 | 3,312 | 625 |

Note: Estimated coefficients and robust standard errors (SE) in parentheses; All models include the same controls as the main analysis; Full results are available upon request.

